# Statistical Analysis Plan for the Stepped-wedge Cluster Randomized Controlled Trial of Electronic Early Notification of Sepsis in Hospitalized Ward Patients (SCREEN) : a study protocol for a stepped-wedge cluster randomized controlled trial

**DOI:** 10.1101/2021.05.23.21257663

**Authors:** Yaseen M Arabi, Ramesh Kumar Vishwakarma, Hasan M Al-Dorzi, Eman Al Qasim, Sheryl Ann Abdukahil, Fawaz K Al-Rabeah, Huda Al Ghamdi, Ebtisam Al Ghamdi, the SCREEN Trial Group

**Author notes:** **Corresponding Author Yaseen Arabi, MD, FCCP, FCCM, ATSF**, College of Medicine, King Saud Bin Abdulaziz University for Health Sciences, King Abdullah International Medical Research Center, Intensive Care Department, King Abdulaziz Medical City, Ministry of National Guard Health Affairs, Riyadh, Saudi Arabia.

## Abstract

**Background:** It is unclear whether screening for sepsis using an electronic alert in hospitalized ward patients improves outcomes. The objective of the SCREEN (Stepped-wedge Cluster Randomized Controlled Trial of Electronic Early Notification of Sepsis in Hospitalized Ward Patients) trial is to evaluate whether an electronic screening for sepsis compared to no screening among hospitalized ward patients reduces all-cause 90-day in-hospital mortality.

**Methods and design:** This study is designed as a stepped-wedge cluster randomized controlled trial in which the unit of randomization or cluster is the hospital ward. An electronic alert for sepsis was developed in the electronic medical record (EMR), with the feature of being active (visible to treating team) or masked (inactive in EMR frontend for the treating team but active in the backend of the EMR). Forty-five clusters in 5 hospitals are randomized into 9 sequences of 5 clusters each to receive the intervention (active alert) over 10 periods, 2-months each, the first being the baseline period. Data are extracted from EMR and are compared between the intervention (active alert) and control group (masked alert). The primary outcome of all-cause hospital mortality by day 90 will be compared using a generalized linear mixed model with a binary distribution and a log-link function to estimate the relative risk as a measure of effect. We will include two levels of random effects to account for nested clustering within wards and periods and two levels of fixed effects; hospitals and COVID-19 ward status. Results will be expressed as relative risk with a 95% confidence interval (CI).

**Conclusion:** The SCREEN trial provides an opportunity for a novel trial design and analysis of routinely collected and entered data to evaluate the effectiveness of an intervention (alert) for a common medical problem (sepsis in ward patients). In this statistical analysis plan, we outline details of the planned analyses in advance of trial completion. Prior specification of the statistical methods and outcomes analysis will facilitate unbiased analyses of these important clinical data.

**Trial registration:** ClinicalTrials.gov NCT04078594. Registered on September 6, 2019, https://clinicaltrials.gov/ct2/show/NCT04078594

## Background

Sepsis is a major cause of morbidity and mortality among hospitalized patients. Sepsis outcome is greatly dependent on the time-sensitive administration of appropriate antimicrobials, fluid resuscitation, and source control.(1-3) Screening for sepsis using an electronic alert in hospitalized patients may improve outcomes by early sepsis recognition and timely implementation of appropriate care processes. However, the evidence for such an intervention is modest,(4) and a randomized controlled trial is needed to measure its true effect.

The objective of the Stepped-wedge Cluster Randomized Controlled Trial of Electronic Early Notification of Sepsis in Hospitalized Ward Patients (SCREEN) is to evaluate whether electronic screening for sepsis compared to no screening among hospitalized ward patients reduces all-cause 90-day in-hospital mortality. The study protocol has been submitted for publication.(5) The electronic screening for sepsis is based on the quick Sequential Organ Failure Assessment qSOFA.(6)

In this manuscript, we describe the statistical analysis plan (SAP) of the SCREEN trial. This SAP complies with the International Conference on Harmonization of Technical Requirements for Registration of Pharmaceuticals for Human Use, and both the “Statistical principles for clinical trials E9” and “Structure and content of clinical study reports E3”.(7, 8) The final study report will follow the CONSORT (Consolidated Standards of Reporting Trials) 2010 guidelines for reporting randomized controlled trials and the CONSORT extension for stepped-wedge cluster randomized trials.(9-11) This SAP identifies the procedures to be applied to the primary and secondary analyses for the trial cohort once trial data are complete. The SAP was finalized during trial implementation, and all analyses were prospectively defined.

## Methods

### Study Design

The study is conducted in the 5 Ministry of National Guard Health Affairs (MNGHA) hospitals which share the same Electronic Medical Record (EMR) system (BESTCare, South Korea). This study is designed as a stepped-wedge cluster randomized controlled trial, which allows to sequentially deliver the study intervention to all trial clusters over a number of periods. We present a glossary of terms in line with the CONSORT extension for stepped-wedge cluster randomized trials(11) in Table 1. The cluster refers to the unit of randomization, which is the hospital ward, and we will refer to it in the rest of the document as “ward”. A list of ward-level and patient-level eligibility criteria is outlined in the study protocol.(5) Wards are randomized into 9 sequences of 5 wards each to receive the intervention. After a baseline period of 2 months, the intervention is implemented in a new sequence of five new randomly selected wards, until it is eventually implemented in all 9 sequences (45 wards) (Figure 1). A computer-generated non-stratified randomly allocated concealed list determines the order in which the wards receive the intervention. The randomization list was maintained with a research coordinator who was not involved in this trial, and the ward allocation remained concealed from the research and clinical teams throughout the study and was revealed for a given sequence only 1 month before the implementation of the intervention to allow training.

**Table 1:**
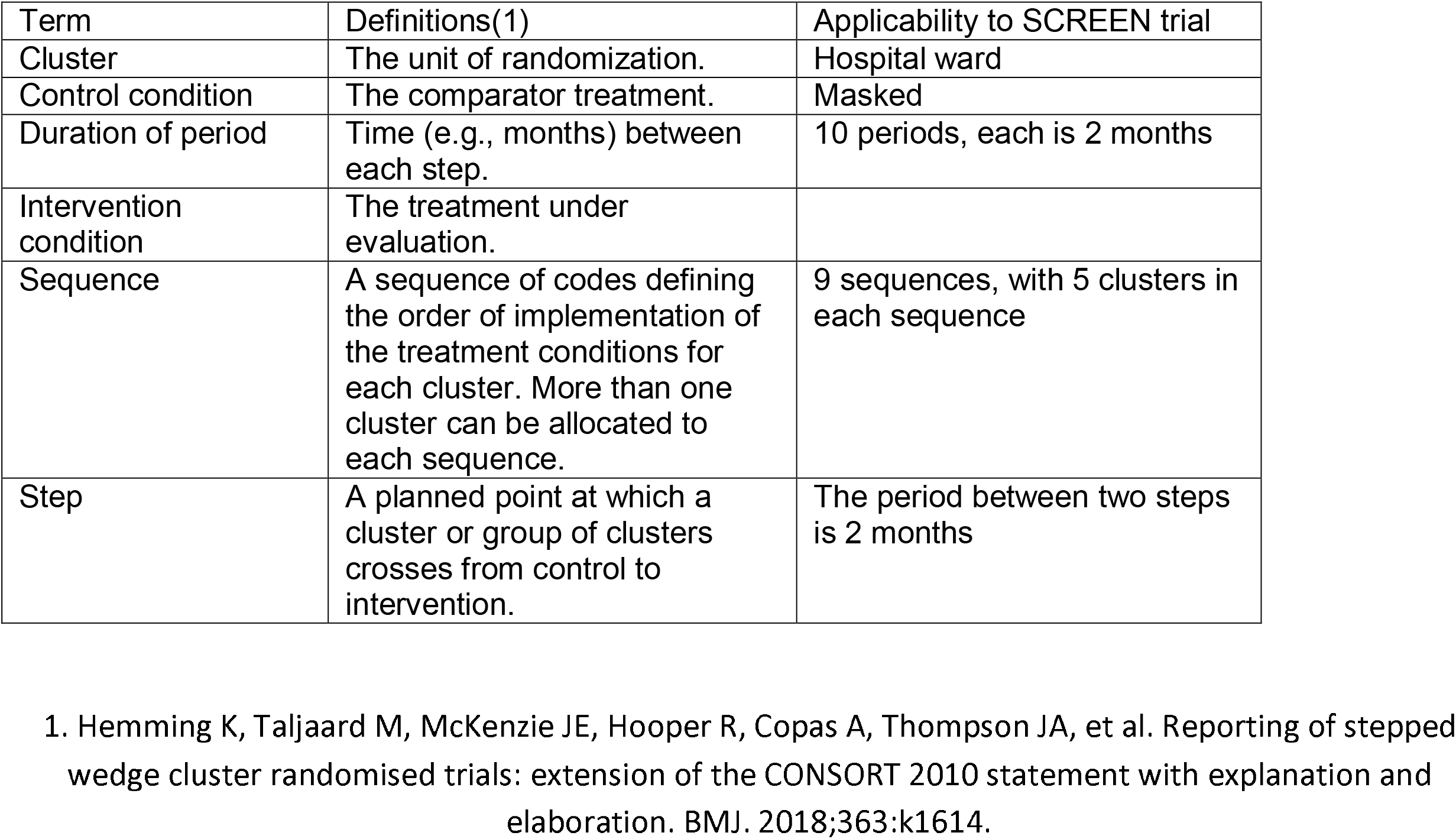
Glossary of terms used in the current report, based on the CONSORT extension for cluster randomized trials(1)

| Term | Definitions(1) | Applicability to SCREEN trial |
| --- | --- | --- |
| Cluster | The unit of randomization. | Hospital ward |
| Control condition | The comparator treatment. | Masked |
| Duration of period | Time (e.g., months) between each step. | 10 periods, each is 2 months |
| Intervention condition | The treatment under evaluation. |  |
| Sequence | A sequence of codes defining the order of implementation of the treatment conditions for each cluster. More than one cluster can be allocated to each sequence. | 9 sequences, with 5 clusters in each sequence |
| Step | A planned point at which a cluster or group of clusters crosses from control to intervention. | The period between two steps is 2 months |

1. Hemming K, Taljaard M, McKenzie JE, Hooper R, Copas A, Thompson JA, et al. Reporting of stepped wedge cluster randomised trials: extension of the CONSORT 2010 statement with explanation and elaboration. *BMJ*. 2018;363:k1614.

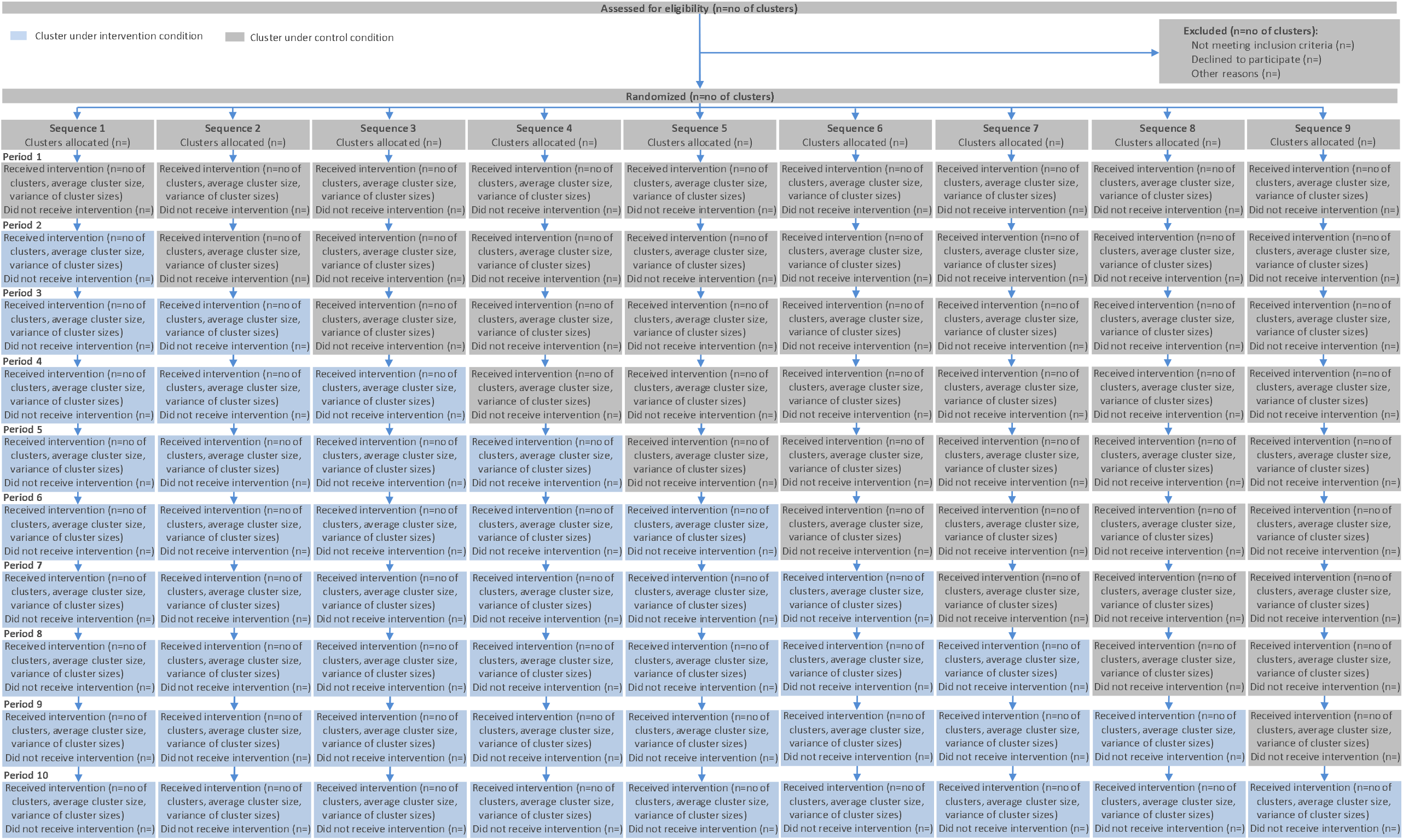

### Intervention and Control Groups

The intervention included the implementation of an electronic alert system and the associated training, feedback and audit. The alert was developed in the electronic medical record (EMR), with the feature of being active (visible to treating team) or masked (inactive in EMR frontend for the treating team but active in the backend of the EMR). Once a ward is randomized to the intervention group, the alert system was activated. The intervention group constitutes of patients admitted to the wards with the active alert, and the control group constitutes of patients admitted to the wards with masked alert.

In the intervention group, once a patient meets the qSOFA criteria, an alert appears in EMR as a pop-message and appears also on a hand-held device carried by the charge nurse of the corresponding ward. The alert prompts the nurse to notify the physician and prompts the physician to assess the patient for possible sepsis. At the beginning of the study, we launched a hospital-wide sepsis awareness campaign in all 5 participating hospitals focusing on the importance of timely interventions for sepsis. We also provided in-service training sessions to the related medical and nursing departments at the beginning of the study and before implementation in each ward. We conducted regular webinars with the leaders of active wards. We also created an intranet page with educational resources (videos, presentations, documents, posters and related links) that explain the project and provides clinical guidance and resources. We developed a dashboard to display data for each active ward on the number of alerts and the percentage and time to acknowledgment by nurses and physicians. We have set 15 minutes as an over-stretch target for nursing acknowledgment and 30 minutes for physician acknowledgment and we have tracked these indicators on the electronic dashboards. The data on dashboards have been made available in real-time for nursing and medical managers of each ward. In addition, reports have been generated every two weeks of the same data and shared with the teams in the active wards. The hospital administration and quality management department has been engaged in the process of feedback and of improving the response time to alerts. The bedside management and assessment have been at the discretion of the treating team, and the project has adopted the 2016 Surviving Sepsis Campaign guidelines and the hour-1 bundle, but without specific monitoring of bundle compliance.(1, 2)

### Study Population

The study has been conducted in the 5 Ministry of National Guard Health Affairs (MNGHA) hospitals: King Abdulaziz Medical City-Riyadh, King Abdulaziz Medical Center-Jeddah, and Prince Mohammed Bin Abdul Aziz Hospital – Al Madinah, King Abdulaziz Hospital - Al Ahsa, and Imam Abdulrahman Al Faisal Hospital – Dammam. A CONSORT diagram will be generated according to the CONSORT extension for stepped-wedge cluster randomized trials (CRTs).(11); for each sequence at each period, the number of clusters receiving the intervention, the average cluster size, and its variance, and the number of clusters not receiving intervention will be presented (Figure 1).

### Sample Size

The sample size for this stepped-wedge cluster-randomized design was calculated for 45 clusters with 10 periods (including one baseline period) using Power Analysis and Sample Size (PASS) software (PASS 15 Power Analysis and Sample Size Software (2017). NCSS, LLC. Kaysville, Utah, USA, ncss.com/software/pass). After a baseline period, 5 new clusters (12) switch from the control group to the intervention group at the beginning of each subsequent period. Using historical data obtained from the development domain of the EMR for ward patients admitted from 01 July 2018 to 30 June 2019, we calculated a baseline in-hospital mortality rate by day 90 of 3.13%. Based on the same dataset, 18.3% of eligible ward patients had an alert based on qSOFA criteria(6) with in-hospital mortality of 8.16% compared to 2% in the non-patients. For sample size calculations, we made the following assumptions: A) the impact of the intervention on mortality occurs only in patients who have the alert, B) Only half of the patients with the alert have sepsis, C) 90% of deaths among the patients with the alert occurred among septic patients, D) early intervention resulting from the alert will reduce the in-hospital mortality by 50%, i.e. from 8.16% to 4.08% in patients with sepsis and would lead to an overall change in in-hospital mortality for the whole cohort from 3.13% to 2.46% yielding a relative risk of 0.79 and a risk difference of 0.67% which is the target effect size, D) 80% power using two-sided Wald Z-Test and significance level of 5% and, E) an intra-cluster correlation (ICC, a measure of the relatedness of cluster) of 0.22 as estimated from the same retrospective electronic database. As the primary analysis would be adjusted for random effect to account for the correlation between patients within the same cluster, we used the estimation variance (σ^2^), which is calculated from responses (P1, P2), as the within-cluster variance (σw^2^) as suggested by Hussey and Hughes (2007) and Hemming and Girling (2014).(13, 14) As such, a reduction of in-mortality in hospital by 90 days from 3.13% to 2.46% (relative risk of 0.79 and a risk difference of 0.67%) requires a total sample size of 65250 subjects (average of 1450 subjects per cluster with an average of 145 subjects per cluster per period). With all five hospitals combined, this is expected to require 20 months (2 months per period). There is no planned interim analysis.

### Study cohorts

#### a. Intention-to-treat cohort

We will report patient flow according to CONSORT flowchart for the stepped-wedge cluster randomized trial by allocated sequence and period.(11)(Figure 1). The intention-to-treat (ITT) cohort includes all eligible patients admitted to the eligible wards. The ITT analysis also implies that patients in the ITT cohort in the wards belonging to a particular period will be analyzed per their planned randomization regardless of what happens during the trial. For example, if a ward was planned to have the intervention during a given period and for technical reasons that alert system was not operational, patients recruited in that ward during that period will be analyzed as receiving an active alert. Although it is not anticipated that there will be wards that crossover their allocated group (i.e., change from alert to non-alert, or vice versa), any such instances will be documented. Patients who are transferred from one ward to another will be counted as part of the first ward. The primary analysis will be based on this population.

The study was launched in October 2019. During the study period in 2020-2021, and in response to the surge in the number of hospitalized patients with coronavirus disease-19 (15), some of the wards had to be converted to intensive care units (16), making them ineligible for the study intervention. These wards will be excluded from the ITT cohort while used as ICUs. During the peak of COVID-19 cases, total admissions to the wards declined substantially to less than 50%, therefore 2 consecutive periods (starting June 2019) were extended from 2 months to 3 months each to account for the decline in cluster size. In addition, some wards were designated for admission of suspected or confirmed COVID-19 cases. Given the higher mortality associated with such designation, and because such designation would likely be over-represented in the intervention group, data on ward designation as a COVID-19 ward will be documented as a ward-level variable, and it will be used as a fixed effect term in the primary model (as discussed below). Additionally, patient-level data on COVID-19 diagnosis will be also obtained.

#### b. Alert cohort

This cohort represents the subset of ITT patients who had the alert whether in the intervention wards or the control wards.

Statistical tests and their confidence intervals (CIs) will be calculated with two-sided. The statistical significance level set will be at the 5% level. All analyses will be performed using SAS version 9.4. Categorical variables will be summarized as counts and frequencies, and continuous variables and median and interquartile ranges or means and standard deviations, if deemed normally distributed.

Reporting Baseline Characteristics, Physiological Parameters and Treatments Baseline characteristics will be presented for the ITT and alert cohorts (Table S1) including age, sex, source of admission, admitting ward, comorbidities (extracted based on ICD-10AM), Charlson comorbidity index, source of infection (pneumonia, urinary tract infection, skin and soft tissue infection, intra-abdominal infection, other infections, and no clear source, extracted based on ICD-10AM) and dialysis. We will also report vital signs (systolic blood pressure, diastolic blood pressure, heart rate, temperature, and respiratory rate) as well as laboratory parameters (lactate, white blood cells, bilirubin, creatinine) and whether culture of blood, respiratory, urine or other body fluids (pleural, ascitic, CSF, joint) were obtained and whether treatments (intravenous fluids and antibiotics) were received at baseline. Definitions of these variables were outlined in the study protocol.(5)

### Reporting Alert Information

We will report the number of patients with at least one alert, and the number of alerts per patient, the time to first alert and the alert information criteria which led to the alert trigger (Table S2). We will report vital signs in the 12 hours pre-alert (systolic blood pressure, diastolic blood pressure, heart rate, temperature, and respiratory rate) as well as laboratory parameters in the 12 hours pre-alert (lactate, white blood cells, bilirubin, creatinine) and whether a culture of blood, respiratory, urine or other body fluids (pleural, ascitic, CSF, joint) were obtained and whether treatments (intravenous fluids and antibiotics) were received in the 12 hours pre-alert.

### Reporting Process Measures and Post-alert Physiologic Parameters

We will compare the following process measures between the two groups: A) percentage of patients with lactate reported within 12 hours of alert if not reported in the 12 hours before alert and highest lactate value reported in the 12 hours after the alert, B) the percentage of patients with blood culture ordered in 12 hours if not performed in the 12 hours before the alert, C) percentage of patients with respiratory, urine and body fluid cultures ordered in 12 hours if not performed in the 12 hours before the alert, D) intravenous fluid administered in 12 hours after alert (yes, no), E) percentage of patients who were not on antibiotics in the 12 hours before alert and had new antibiotic administered within 3 and 12 hours of the alert, F) post-alert systolic and diastolic blood pressure: (lowest value in the 12 hours after the alert) and heart and respiratory rate (highest value in the 12 hours after the alert) (Table S2).

### Study Outcomes

The primary outcome is defined as all-cause in-hospital mortality within 90 days. Other outcomes include hospital length of stay (LOS) (ITT cohort and alert cohort), transfer to ICU within 90 days (ITT cohort and alert cohort) and 14 days of alert (alert cohort), ICU-free days in the first 90 days (ITT cohort and alert cohort), critical care response team (CCRT) activation within 90 days (ITT cohort and alert cohort) and 14 days of alert (alert cohort), cardiac arrest within 90 days (ITT cohort and alert cohort) and 14 days of alert (alert cohort), the need for mechanical ventilation, vasopressor therapy and incident renal replacement therapy within 90 days (all in ITT cohort and alert cohort) and 14 days of alert (alert cohort). Balancing/ safety outcomes measures include antibiotic-free days, acquisition of multidrug-resistant organisms within 90 days in both groups (ITT cohort and alert cohort) and acquisition of Clostridium difficile infection within 90 days (ITT cohort and alert cohort) (Table S3 and S4).

### Analysis of Primary Outcome

The primary outcome of all-cause in-hospital mortality by day 90 will be compared between the intervention group and control group at the individual level with the use of mixed-effect logistic regression model with jack-knife method to estimates standard errors to account for grouping within clusters.(13, 17) We will use a generalized linear mixed model with a binary distribution and a log-link function to estimate the relative risk as a measure of effect. We will include two levels of random effects to account for nested clustering within wards and periods and two levels of fixed effects; hospitals and COVID-19 ward status. The model will be selected as the best model with a unique covariance structure that produces the lowest Bayesian Information Criterion (BIC) value. The covariance structures that will be considered in the model are: first order of autocorrelation covariance structure, unstructured covariance structure, Toeplitz covariance structure and variance component structure (VC). We will use Satterthwaite method to adjust for denominator degree of freedom for tests of the fixed effects. The random coefficients will be modeled using G-side random effects and obtain the subject-specific estimates by defining the appropriate variance-covariance structure. The overdispersion of parameters will also be assessed by calculating the ratio of the Pearson chi-square statistic and its degrees of freedom. If this ratio exceeds 1, it indicates that the variability in data has not been properly modeled and that there is residual overdispersion due to misspecification of the conditional distribution. In case of no event reported in a cluster for a particular period, a firth correction will be used in the generalized linear mixed model to avoid the issue of quasi-separation. The Newton-Raphson optimization technique with ridging option might be used to help with the convergence of the model. Results will be expressed as relative risk with 95% confidence interval (CI). The primary outcome will be also analyzed similarly in the alert cohort. In case any models fail to converge, we will report ORs from mixed logistic regression. We will also estimate risk difference.(18)

### Analysis of secondary outcomes

Categorical outcomes including ICU admission, CCRT activation, cardiac arrest, the need for mechanical ventilation, vasopressor therapy, incident renal replacement therapy, acquisition of multidrug-resistant organisms and Clostridium difficile infection will be compared between the intervention and the control group, in a similar model to the one used in the analysis primary outcome. We will include two levels of random effects to account for nested clustering within wards and periods and two levels of fixed effects; hospitals and COVID-19 ward status. Results will be expressed as relative risk with 95% CI.

Continuous outcomes including hospital length of stay (LOS), ICU-free days, and antibiotic-free days will be compared using a mixed effect Poisson model (Table S3). We will include two levels of random effects to account for nested clustering within wards and periods and two levels of fixed effects; hospitals and COVID-19 ward status. The model will be selected as best model with a unique covariance structure that produces the lowest Bayesian Information Criterion (BIC) value. The covariance structures that will be considered in the model are: first order of autocorrelation covariance structure, unstructured covariance structure, Toeplitz covariance structure and variance component structure (VC).” The random coefficients will be modeled using G-side random effects and obtain the subject-specific estimates by defining the appropriate variance-covariance structure. The results will be expressed as beta estimates with 95% CI.

### Sensitivity and subgroup analyses

To address the concern about contamination, we will conduct a sensitivity analysis excluding all patients in the control group in the 90 days before crossing over to the intervention group. Because there are fewer than 50 clusters, we will conduct a sensitivity analysis using a small sample correction with the Kenward-Roger method.(13, 19, 20) We will conduct a sensitivity analysis adjusting for the following covariates: type of wards (medical, surgical, oncology and mixed), age, baseline systolic blood pressure, baseline respiratory rate, GCS and Charlson comorbidity index. For the latter analysis, we will use imputation for missing variables as outlined below. In addition, we will conduct also a complete case sensitivity analysis. We will conduct a sensitivity analysis excluding the periods in which wards were assigned as COVID-19 ward.

We will analyze the primary outcome of all-cause hospital mortality by day 90 across predefined subgroups using the same model of the primary analysis. The predefined subgroups include age=< 65 years and >65 years, patients with documented infection source (including ICD-10AU for pneumonia, urinary tract infection, skin and soft tissue infection, intra-abdominal infection or other infections) and patients with no documented infection or infection source, patients admitted to medical, surgical, oncology and mixed wards, alert within 48 hours of admission and after 48 hours of admission and patients admitted to COVID-19 and non-COVID-19 wards. (Table S6). Results of the test of interaction will be reported.

### Handling dropouts and missing data

We do not expect to have missing observations in the variables required for the primary analysis. Given the nature of the study, missing observations are expected, for example, not all patients will have all laboratory tests and therapies within the narrow time windows defined in the protocol. Variables used in the descriptive analysis will not be imputed. In some of the secondary and sensitivity analyses, imputation will be used. Because Glasgow outcome score is not documented for patients with normal neurologic status, missing observations will be assigned a normal value for purposes of the model adjustment. Other variables used in the model adjustment model will be assessed and characterized in terms of their pattern (i.e., Missing Completely at Random, Missing at Random, Missing Not at Random). For Missing Completely at Random data, all analyses will be based on a list-wise deletion approach where observation will complete values will be only considered for analysis. For variables with values Missing at Random, multiple imputation techniques will be utilized to impute the missing values as suggested by Rubin.(21) And for variables with values Missing Not at Random, a pattern-mixture model technique will be used to impute the missing values.(22)

### Graphical Presentation

The subgroup analyses will be displayed as a forest plot.

### Adjustment for multiplicity

To adjust for multiple testing for secondary outcomes and subgroup analyses, we will use the False Discovery Rate (FDR) as described by Benjamini and Hochberg.(23) In this procedure all hypothesis tests will be sorted in descending order based on their calculated p-value. All hypothesis tests below an index K will be rejected where K is calculated as follows: 
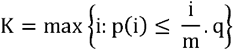

where i =m, …, 1, m is the total number of tested hypotheses; q = 0.05. The multiplicity testing adjustment will also be done on confidence intervals by constructing 1-K*q/m CI for each selected parameter.(12)

### Protocol deviations

We will report protocol deviations, if they occur, including failure to implement the intervention in a given ward, or wrong implementation of the intervention in a ward assigned to the control group. These deviations will be documented on the CONSORT flow diagram. The data from such wards will be analyzed according to the ITT principle.

### Study governance

The study management and development committee is responsible for the overall management of the study, data management and maintenance of the trial master file and statistical master file following the Good Clinical Practice principles.

## Discussion

The SCREEN trial provides an opportunity for a novel trial design and analysis of routinely collected and entered data to evaluate the effectiveness of an intervention for a common medical problem (sepsis in ward patients).

Stepped-wedge cluster randomized trials involve randomization of clusters to different sequences, and are increasingly used in clinical medicine.(14, 24-26) This design is suitable for evaluating interventions delivered at the level of the cluster. In the SCREEN trial, it allows the assessment of the effect of the alert system by comparing the outcomes of patients in the intervention and control cohorts as well as over time.(11) In addition, by having the masked alert, an additional comparison between patients with alerts in the intervention and control groups will be performed.

Strengths of our statistical analysis plan include the analysis according to the intention to treat principle, the large sample size and the multicenter nature. Our analysis plan highlights some potential trial limitations, in relation to the low event rate, which we have addressed by the large sample size. We will address the multiplicity of secondary analyses by reporting the False Discovery Rate. Our alert which is based on qSOFA is possibly applicable in other healthcare settings, although there may be variations across different EMRs. As per the design of the stepped-wedge trial, the proportion of clusters in the intervention group increases gradually with time; and as a result, the intervention group will, on average, have more observations at later dates of the study.(14) Because external factors may influence outcome over time, the time is a potential confounder and was accounted in the sample size calculation and will be adjusted for in the analysis. (14) COVID-19 pandemic is an example of an external factor that is associated with increased in-hospital mortality of hospitalized patients.(27) Because certain wards were assigned as intensive care units during the pandemic, we excluded these wards for that duration of ICU assignment from the analysis. Other wards were assigned as COVID-19 wards. To address this confounder, we will account for COVID-19 ward status as a fixed effect term for each period. In addition, we will carry out a sensitivity analysis excluding the periods in which wards were assigned as COVID-19 ward. We will also conduct a subgroup analysis by whether patients were admitted to COVID-19 or non-COVID-19 wards. We used 90-day in-hospital mortality rather than a shorter mortality measure as the primary outcome to capture the effect of the intervention on a longer outcome. However, this would likely cause some degree of within-cluster contamination, as some patients in the control wards will become exposed to the alert after crossing over to the intervention group. The impact of the contamination is likely to be modest because the average stay in the hospital is likely to be <10 days. Nevertheless, we will conduct a sensitivity analysis excluding all patients in the control group in the 90 days before crossing over to the intervention group. We did not stratify randomization according to hospital given that the 5 hospitals follow similar healthcare delivery model; however, we used hospital as a fixed effect term in the analytical model.

The study has started in October 2019 and is anticipated to have complete implementation and follow-up data by the end of October 2021.

## Conclusion

In this statistical analysis plan, we outline details of the planned analyses in advance of SCREEN trial completion. Prior specification of the statistical methods and outcomes analysis will facilitate unbiased analyses of these important clinical data.

## Data Availability

The datasets will be available from the corresponding author as per the regulations of King Abdullah International Medical Research Center (KAIMRC).

## Abbreviations

CCRT: Critical Care Response Team
CI: Confidence interval
EMR: Electronic Medical Record
ICU: Intensive Care Unit
ITT: Intention to Treat
LOS: Length of Stay
qSOFA: Quick Sequential Organ Failure Assessment
SOFA: Sequential Organ Failure Assessment

## Declarations

### Ethics approval and consent to participate

The study was approved by the Institutional Review Board of the Ministry of National Guard - Health Affairs. Informed consent was waived because of the nature of the study.

### Consent for publication

Not applicable.

### Authors’ contributions

YA is the Chief Investigator; conception and design, the analytical plan, drafting of the manuscript, critical revision of the manuscript for important and intellectual content. RV, HD, EQ, SA, FR, HG, EG contributed to the development of the protocol, critical revision of the manuscript for important intellectual content. All authors agree to be accountable for all aspects of the work and have read and approved the final manuscript.

## Acknowledgements

We would like to thank Dr. Jessica Kasza, Associate Professor in biostatistics in the School of Public Health and Preventive Medicine at Monash University, for providing statistical input to this SAP.

**Collaborators: the SCREEN Trial Group\***

**Management and Development Group:**

**Yaseen M Arabi, MD, FCCP, FCCM, ATSF**

Chairman, Intensive Care Department

Medical Director, Respiratory Services

King Abdulaziz Medical City, Central Region

Professor, College of Medicine,

King Saud Bin Abdulaziz University for Health Sciences

Ministry of National Guard Health Affairs, Riyadh, Saudi Arabia

**Abdulmohsen Alsaawi, MD, MSc PH, MSc EBHC**

Executive Director, Medical Services

Consultant, Emergency Medicine Department

King Abdulaziz Medical City, Central Region

Assistant Professor, College of Medicine,

King Saud Bin Abdulaziz University for Health Sciences

Ministry of National Guard Health Affairs, Riyadh, Saudi Arabia

**Mohammed Al Zahrani, FACS, MPH, SSC-Surg**

Executive Director, Medical Services

Consultant, Surgical Oncology

King Abdulaziz Medical City, Western Region

Ministry of National Guard Health Affairs, Jeddah, Saudi Arabia

**Ali Al Khathaami, MD, MPH, FRCPC**

Executive Director, Quality and Patient Safety Department

King Abdulaziz Medical City, Central Region

Ministry of National Guard Health Affairs, Riyadh, Saudi Arabia

**Raed H. AlHazme, PhD**

Executive Director, Information Systems and Informatics Division (ISID)

King Abdulaziz Medical City, Central Region

Ministry of National Guard Health Affairs, Riyadh, Saudi Arabia

**Abdullah Al Mutrafy, MD**

Deputy Executive Director, Medical Services

King Abdullah Specialized Children’s Hospital

Ministry of National Guard Health Affairs, Riyadh, Saudi Arabia

**Ali Al Qarni, MD, SBIM, JBIM, FACP, FRCP, FACE**

Deputy Executive Regional Director, Medical Services, Al Ahsa

Ministry of National Guard Health Affairs, Al Ahsa, Saudi Arabia

**Ahmed Al Shouabi, MD SBFM, ABFM, MSc**

Deputy Exectuive Regional Director, Medical Services,

Dammam Imam Abdulrahman Al Faisal Hospital

Ministry of National Guard Health Affairs, Dammam, Saudi Arabia

**Amar Alhasani, MD**

Deputy Executive Director-Medical services

Prince Mohammed bin Abdulaziz Hospital

Ministry of National Guard Health Affairs, Madinah, Saudi Arabia

**Eman Al Qasim, RN, MSN**

Clinical Research Coordinator II, Intensive Care Department

Research Office, King Abdullah International Medical Research Center

King Saud Bin Abdulaziz University for Health Sciences

King Abdulaziz Medical City, Central Region

Ministry of National Guard Health Affairs, Riyadh, Saudi Arabia

**Sheryl Ann Abdukahil, RN**

Quality Management Specialist II, Intensive Care Department

King Abdulaziz Medical City, Central Region

Ministry of National Guard Health Affairs, Riyadh, Saudi Arabia

**Fawaz K Al-Rabeah, RPh, MHI**

Manager, Medical Application, Corporate Clinical Information Management System

Information Systems and Informatics Division

King Abdulaziz Medical City, Central Region

Ministry of National Guard Health Affairs, Riyadh, Saudi Arabia

**Huda Al Ghamdi**

Manager, Applications, Department of Database Information Management,

Information Systems and Informatics Division

King Abdulaziz Medical City, Central Region

Ministry of National Guard Health Affairs, Riyadh, Saudi Arabia

**Ebtisam Al Ghamdi**

Application Analyst, Knowledge Management Information Systems and Informatics

Division King Abdulaziz Medical City, Central Region

Ministry of National Guard Health Affairs, Riyadh, Saudi Arabia

**Abdulaleem Alattasi, MBBS, FRCPC**

Consultant, Anesthesia & ICU

Deputy Chairman, Department of Pediatric Anesthesia – KASCH

Director, Faculty Development Program, KSAU-HS

Director of Operating Room Services, KASCH

Deputy Executive Director, Perioperative Quality and Patient Safety

King Abdulaziz Medical City, Central Region

Ministry of National Guard Health Affairs, Riyadh, Saudi Arabia

**Raed Al Almoodi**

Programmer Analysit ISID EMS HIS

**Hasan M. Al-Dorzi, MD**

Section Head, Trauma ICU & Medical ICU

Consultant, Intensive Care Department

King Abdulaziz Medical City, Central Region

Ministry of National Guard Health Affairs, Riyadh, Saudi Arabia

**Ramesh Kumar Vishwakarma, PhD**

Biostatistician, Section of Biostatistics

King Abdullah International Medical Research Center

King Saud Bin Abdulaziz University for Health Sciences

Ministry of National Guard Health Affairs, Riyadh, Saudi Arabia

**Implementation Riyadh:**

**Khadega A. Abuelgasim, MD**

Consultant, Adult Hematology & Oncology Department

King Abdulaziz Medical City, Central Region

Ministry of National Guard Health Affairs, Riyadh, Saudi Arabia

**John Alchin, RN, MHA**

Director, Clinical Nursing, Critical Care, Nursing Services Department,

King Abdulaziz Medical City, Central Region

Ministry of National Guard Health Affairs, Riyadh, Saudi Arabia

**Ahmad Alharbi, MBBS**

Consultant, Infectious Diseases, Medicine Department

King Abdulaziz Medical City, Central Region

Ministry of National Guard Health Affairs, Riyadh, Saudi Arabia

**Mufareh Edah AlKatheri, MD, MBBS**

Director, Quality and Patient Safety Department

King Abdulaziz Medical City

Ministry of National Guard Health Affairs, Riyadh, Saudi Arabia

**Joan Jones, RN, MA (Health Management)**

Nurse Manager, Medical-Nursing Services Department

King Abdulaziz Medical City, Central Region

Ministry of National Guard Health Affairs, Riyadh, Saudi Arabia

**Saad Al-Qahtani, MD, MMED, MAHA, FRCPC**

Section Head, Critical Care Response Team

Chairman, Hospital Mortality & Morbidity Committee

Consultant, Intensive Care Department

Associate Professor, College of Medicine,

King Saud Bin Abdulaziz University for Health Sciences

King Abdulaziz Medical City, Central Region

Ministry of National Guard Health Affairs, Riyadh, Saudi Arabia

**Salih Bin Salih, MD, FACP, FRCP (Edin)**

Professor, College of Medicine, King Saud Bin Abdulaziz University for Health Sciences

Chairman, Department of Medicine, King Abdulaziz Medical City

Ministry of National Guard Health Affairs, Riyadh, Saudi Arabia

**Nahar Alselaim, MBBS, MPH, FRCSC**

Consultant, Surgery Department

Assistant Professor, College of Medicine, King Saud Bin Abdulaziz University for Health Sciences

King Abdulaziz Medical City

Ministry of National Guard Health Affairs, Riyadh, Saudi Arabia

**Nabiha Tashkandi, BSN, RN, MSN**

Associate Executive Director, Nursing Services

King Abdulaziz Medical City, Central Region

Ministry of National Guard Health Affairs, Riyadh, Saudi Arabia

**Zeyad Alyousef, MD, SSC (Ortho), ABOS**

Chairman, Surgery Department

Assistant Professor, College of Medicine, King Saud Bin Abdulaziz University for Health Sciences

King Abdulaziz Medical City, Central Region

Ministry of National Guard Health Affairs, Riyadh, Saudi Arabia

**Amal Matroud, RN**

Nursing Services Department, King Abdulaziz

Medical City Ministry of National Guard Health Affairs, Riyadh, Saudi Arabia

**Rasha Ebeid Al Anazi, RN**

Nursing Services Department, King Abdulaziz Medical City

Ministry of National Guard Health Affairs, Riyadh, Saudi Arabia

**Implementation Jeddah:**

**Fahad Al-Hameed, MD, FRCPC**

Chairman, Intensive Care Department,

King Abdulaziz Medical City, Western Region

Ministry of National Guard Health Affairs, Jeddah, Saudi Arabia

**Wasil Jastaniah, MBBS, FAAP, FRCPC**

Professor & Consultant

Pediatric Hematology, Oncology, BMT

Chairman, Princess Noorah Oncology Center

King Abdulaziz Medical City, Western Region

Ministry of National Guard Health Affairs, Jeddah, Saudi Arabia

**Hassan Ahmad AlMarhabi, MD**

Director, Quality and Patient Safety Department

Assistant Professor, Internal Medicine & Infectious Diseases

College of Medicine, Jeddah, King Saud Bin Abdulaziz University for Health Sciences

Consultant Adult & Transplant Infectious Diseases, Internal Medicine

King Abdulaziz Medical City, Western Region

Ministry of National Guard Health Affairs, Jeddah, Saudi Arabia

**Emad AlWafi, MD**

Section Head, Internal Medicine

King Abdulaziz Medical City, Western Region

Ministry of National Guard Health Affairs, Jeddah, Saudi Arabia

**Ali H. Alyami, MD**

Chairman, Surgery Department

King Abdulaziz Medical City, Western Region

Ministry of National Guard Health Affairs, Jeddah, Saudi Arabia

**Arwa O Yamani, MD**

Deputy Chairman Oncology Quality & Patient Safety

King Abdulaziz Medical City, Western Region

Ministry of National Guard Health Affairs, Jeddah, Saudi Arabia

**Mayadah Habshi, BSN. DNE.EFQM.MSN**

Quality Improvement Specialist I

King Abdulaziz Medical City, Western Region

Ministry of National Guard Health Affairs, Jeddah, Saudi Arabia

**Basem Banat, RN**

Manager Nursing Resource Systems

King Abdulaziz Medical City, Western Region

Ministry of National Guard Health Affairs, Jeddah, Saudi Arabia

**Omar Abuskout, RN**

Director Clinical Nursing

King Abdulaziz Medical City, Western Region

Ministry of National Guard Health Affairs, Jeddah, Saudi Arabia

**Adnan Munshi, Eng**

Manager Picture Archiving & Communication System

King Abdulaziz Medical City, Western Region

Ministry of National Guard Health Affairs, Jeddah, Saudi Arabia

**Implementation: Alhasa**

**Hani T. Mustafa, FRACP**

Chairman, Department of Medicine, King Abdulaziz Hospital

Ministry of National Guard Health Affairs, Al Ahsa, Saudi Arabia

**<u>Sami Musalam Aliyyen, RN</u>**

Quality & Patients Safety Department

King Abdulaziz Hospital

Ministry of National Guard Health Affairs, Al Ahsa, Saudi Arabia

**Christa Myumi Sian, RN**

Clinical Resource Nurse King Abdulaziz Hospital

Ministry of National Guard Health Affairs, Al Ahsa, Saudi Arabia

**Abdulaziz Al Qasem**

Information Systems and Informatics Division, King Abdulaziz Hospital

Ministry of National Guard Health Affairs, Al Ahsa, Saudi Arabia

**Implementation Dammam:**

**Gaber Madram, MD**

Consultant Internal Medicine,

Imam Abdulrahman Al Faisal Hospital

Ministry of National Guard Health Affairs, Dammam, Saudi Arabia

**Wafa Nasser, MD**

Director Infection Prevention & Control Program, Infection Control

Imam Abdulrahman Al Faisal Hospital

Ministry of National Guard Health Affairs, Dammam, Saudi Arabia

**Shaher Qahtani, MD**

Director, Quality and Patient Safety Department,

Imam Abdulrahman Al Faisal Hospital

Ministry of National Guard Health Affairs, Dammam, Saudi Arabia

**Clara Masala, RN**

Nurse Manager, Imam Abdulrahman Al Faisal Hospital

Ministry of National Guard Health Affairs, Dammam, Saudi Arabia

**Hannan Al Somali, RN**

Manager Nursing resource systems

Imam Abdulrahman Al Faisal Hospital

Ministry of National Guard Health Affairs, Dammam, Saudi Arabia

**Fatimah Talaqof, RN**

Nurse Manager, Imam Abdulrahman Al Faisal Hospital

Ministry of National Guard Health Affairs, Dammam, Saudi Arabia

**Maryam Almulhim**

Information Services Department-Al Dammam, Imam Abdulrahman Al Faisal Hospital

Ministry of National Guard Health Affairs, Dammam, Saudi Arabia

**Implementation Madinah:**

**Ahmad S. Qureshi, MD**

Chairman, Intensive Care Unit

Consultant, Pulmonology-ICU,

Intensive Care Department, Prince Mohammed bin Abdulaziz Hospital

Ministry of National Guard Health Affairs, Madinah, Saudi Arabia

**Mohammad Abd Rabo, MD**

Manager, Quality & patient safety department, Prince Mohammed bin Abdulaziz Hospital

Ministry of National Guard Health Affairs, Madinah, Saudi Arabia

**Hattan Esilan, MD**

Director, Quality and Patient Safety Department

Prince Mohammed bin Abdulaziz Hospital

Ministry of National Guard Health Affairs, Madinah, Saudi Arabia

**Azurahazri Abd Rahim, RN**

Manager, Nursing Department

Prince Mohammed bin Abdulaziz Hospital

Ministry of National Guard Health Affairs, Madinah, Saudi Arabia

**Naif Almughamisi**

Information Services Department, Prince Mohammed bin Abdulaziz Hospital

Ministry of National Guard Health Affairs, Madinah, Saudi Arabia

## REFERENCES

1. Rhodes A, Evans LE, Alhazzani W, Levy MM, Antonelli M, Ferrer R, et al. Surviving sepsis campaign: international guidelines for management of sepsis and septic shock: 2016. Intensive care medicine. 2017;43(3):304–77.

2. Levy MM, Evans LE, Rhodes A. The surviving sepsis campaign bundle: 2018 update. Intensive care medicine. 2018;44(6):925–8.

3. Rivers E, Nguyen B, Havstad S, Ressler J, Muzzin A, Knoblich B, et al. Early goal-directed therapy in the treatment of severe sepsis and septic shock. New England Journal of Medicine. 2001;345(19):1368–77.

4. Makam AN, Nguyen OK, Auerbach AD. Diagnostic accuracy and effectiveness of automated electronic sepsis alert systems: a systematic review. Journal of hospital medicine. 2015;10(6):396–402.

5. Arabi YM, Saawi AA, Zahrani MA, Khathaami AA, AlHazme RH, Mutrafy AA, et al. Electronic early notification of sepsis in hospitalized ward patients: a study protocol for a stepped-wedge cluster randomized controlled trial. medRxiv. 2021:2021.05.20.21257511.

6. Singer M, Deutschman CS, Seymour CW, Shankar-Hari M, Annane D, Bauer M, et al. The third international consensus definitions for sepsis and septic shock (Sepsis-3). Jama. 2016;315(8):801–10.

7. Kahan BC, Harhay MO. Many multicenter trials had few events per center, requiring analysis via random-effects models or GEEs. Journal of clinical epidemiology. 2015;68(12):1504–11.

8. Alexiou VG, Ierodiakonou V, Dimopoulos G, Falagas ME. Impact of patient position on the incidence of ventilator-associated pneumonia: a meta-analysis of randomized controlled trials. Journal of critical care. 2009;24(4):515–22.

9. Moher D, Hopewell S, Schulz KF, Montori V, Gotzsche PC, Devereaux PJ, et al. CONSORT 2010 explanation and elaboration: updated guidelines for reporting parallel group randomised trials. BMJ. 2010;340:c869.

10. Schulz KF, Altman DG, Moher D, Group C. CONSORT 2010 statement: updated guidelines for reporting parallel group randomised trials. BMJ. 2010;340:c332.

11. Hemming K, Taljaard M, McKenzie JE, Hooper R, Copas A, Thompson JA, et al. Reporting of stepped wedge cluster randomised trials: extension of the CONSORT 2010 statement with explanation and elaboration. BMJ. 2018;363:k1614.

12. Benjamini Y, Yekutieli D, Edwards D, Shaffer JP, Tamhane AC, Westfall PH, et al. False Discovery Rate: Adjusted Multiple Confidence Intervals for Selected Parameters [with Comments, Rejoinder]. Journal of the American Statistical Association. 2005;100(469):71–93.

13. Hussey MA, Hughes JP. Design and analysis of stepped wedge cluster randomized trials. Contemp Clin Trials. 2007;28(2):182–91.

14. Hemming K, Haines TP, Chilton PJ, Girling AJ, Lilford RJ. The stepped wedge cluster randomised trial: rationale, design, analysis, and reporting. BMJ. 2015;350:h391.

15. Richardson S, Hirsch JS, Narasimhan M, Crawford JM, McGinn T, Davidson KW, et al. Presenting Characteristics, Comorbidities, and Outcomes Among 5700 Patients Hospitalized With COVID-19 in the New York City Area. Jama. 2020;323(20):2052–9.

16. Borba MGS, Val FdA, Sampaio VS, Alexandre MAA, amp, uacutejo, et al. Chloroquine diphosphate in two different dosages as adjunctive therapy of hospitalized patients with severe respiratory syndrome in the context of coronavirus (SARS-CoV-2) infection: Preliminary safety results of a randomized, double-blinded, phase IIb clinical trial (CloroCovid-19 Study). medRxiv. 2020:2020.04.07.20056424.

17. Shakeshaft A, Doran C, Petrie D, Breen C, Havard A, Abudeen A, et al. The effectiveness of community action in reducing risky alcohol consumption and harm: a cluster randomised controlled trial. PLoS Med. 2014;11(3):e1001617.

18. . McGowan L, Goodman M, https://www.lexjansencom/nesug/nesug13/118_Final_Paperpdf Accessed on Aug 15-2021.

19. Thompson JA, Hemming K, Forbes A, Fielding K, Hayes R. Comparison of small-sample standard-error corrections for generalised estimating equations in stepped wedge cluster randomised trials with a binary outcome: A simulation study. Stat Methods Med Res. 2021;30(2):425–39.

20. Kenward MG, Roger JH. Small sample inference for fixed effects from restricted maximum likelihood. Biometrics. 1997;53(3):983–97.

21. .Little, RJA, and D B Rubin (1987) Statistical Analysis with Missing Data New York: John Wiley & Sons.

22. Thijs H, Molenberghs G, Michiels B, Verbeke G, Curran D. Strategies to fit pattern-mixture models. Biostatistics. 2002;3(2):245–65.

23. Benjamini Y, Hochberg Y. Controlling the false discovery rate: a practical and powerful approach to multiple testing. Journal of the Royal statistical society: series B (Methodological). 1995;57(1):289–300.

24. Beard E, Lewis JJ, Copas A, Davey C, Osrin D, Baio G, et al. Stepped wedge randomised controlled trials: systematic review of studies published between 2010 and 2014. Trials. 2015;16:353.

25. Lloyd M, Karahalios A, Janus E, Skinner EH, Haines T, De Silva A, et al. Effectiveness of a Bundled Intervention Including Adjunctive Corticosteroids on Outcomes of Hospitalized Patients With Community-Acquired Pneumonia: A Stepped-Wedge Randomized Clinical Trial. JAMA Intern Med. 2019;179(8):1052–60.

26. Mdege ND, Man MS, Taylor Nee Brown CA, Torgerson DJ. Systematic review of stepped wedge cluster randomized trials shows that design is particularly used to evaluate interventions during routine implementation. J Clin Epidemiol. 2011;64(9):936–48.

27. Kadri SS, Sun J, Lawandi A, Strich JR, Busch LM, Keller M, et al. Association Between Caseload Surge and COVID-19 Survival in 558 U.S. Hospitals, March to August 2020. Ann Intern Med. 2021.

