## Supplementary file for "Statistical Analysis Plan for the Stepped-wedge Cluster Randomized Controlled Trial of Electronic Early Notification of Sepsis in Hospitalized Ward Patients (SCREEN) : a study protocol for a stepped-wedge cluster randomized controlled trial"

**Table S1:** Baseline data.

|  | **Intention-to-treat cohort** | | **Alert cohort** | |
| --- | --- | --- | --- | --- |
|  | **Intervention (XX)** | **Control**  **(XX)** | **Alert**  **(XX)** | **Control**  **(XX)** |
| Age (yr), Median (Q1, Q3) | xx (xx, xx) | xx (xx, xx) | xx (xx, xx) | xx (xx, xx) |
| Female sex – n (%) | xx (xx) | xx (xx) | xx (xx) | xx (xx) |
| Admission source – n (%) |  |  |  |  |
| Emergency Room | xx (xx) | xx (xx) | xx (xx) | xx (xx) |
| Operating Room | xx (xx) | xx (xx) | xx (xx) | xx (xx) |
| Clinic | xx (xx) | xx (xx) | xx (xx) | xx (xx) |
| Intensive care unit | xx (xx) | xx (xx) | xx (xx) | xx (xx) |
| Others | xx (xx) | xx (xx) | xx (xx) | xx (xx) |
| Admitting ward – n (%) |  |  |  |  |
| Medical | xx (xx) | xx (xx) | xx (xx) | xx (xx) |
| Surgical | xx (xx) | xx (xx) | xx (xx) | xx (xx) |
| Oncology | xx (xx) | xx (xx) | xx (xx) | xx (xx) |
| Mixed (any combination) | xx (xx) | xx (xx) | xx (xx) | xx (xx) |
| Comorbidities – n (%) |  |  |  |  |
| End-Stage Renal Disease (ESRD) | xx (xx) | xx (xx) | xx (xx) | xx (xx) |
| Chronic liver disease | xx (xx) | xx (xx) | xx (xx) | xx (xx) |
| Cancer without metastasis | xx (xx) | xx (xx) | xx (xx) | xx (xx) |
| Immune-compromised | xx (xx) | xx (xx) | xx (xx) | xx (xx) |
| Diabetes non-complicated | xx (xx) | xx (xx) | xx (xx) | xx (xx) |
| Diabetes, complicated | xx (xx) | xx (xx) | xx (xx) | xx (xx) |
| Congestive heart failure | xx (xx) | xx (xx) | xx (xx) | xx (xx) |
| Acquired immunodeficiency syndrome | xx (xx) | xx (xx) | xx (xx) | xx (xx) |
| Moderate to severe chronic kidney disease (CKD) | xx (xx) | xx (xx) | xx (xx) | xx (xx) |
| Myocardial infarction | xx (xx) | xx (xx) | xx (xx) | xx (xx) |
| Chronic pulmonary disease | xx (xx) | xx (xx) | xx (xx) | xx (xx) |
| Peripheral vascular disease | xx (xx) | xx (xx) | xx (xx) | xx (xx) |
| Stroke or transient ischemic attack | xx (xx) | xx (xx) | xx (xx) | xx (xx) |
| Dementia | xx (xx) | xx (xx) | xx (xx) | xx (xx) |
| Hemiplegia or paraplegia | xx (xx) | xx (xx) | xx (xx) | xx (xx) |
| Connective tissue disease | xx (xx) | xx (xx) | xx (xx) | xx (xx) |
| Peptic ulcer disease | xx (xx) | xx (xx) | xx (xx) | xx (xx) |
| Mild liver disease | xx (xx) | xx (xx) | xx (xx) | xx (xx) |
| Moderate to severe liver disease | xx (xx) | xx (xx) | xx (xx) | xx (xx) |
| Charlson Comorbidity Index | xx (xx) | xx (xx) | xx (xx) | xx (xx) |
| Source of infection on admission – n (%) |  |  |  |  |
| Pneumonia | xx (xx) | xx (xx) | xx (xx) | xx (xx) |
| Urinary tract infection | xx (xx) | xx (xx) | xx (xx) | xx (xx) |
| Skin and soft tissue infection | xx (xx) | xx (xx) | xx (xx) | xx (xx) |
| Intra-abdominal infection | xx (xx) | xx (xx) | xx (xx) | xx (xx) |
| Other infections | xx (xx) | xx (xx) | xx (xx) | xx (xx) |
| No documented infection or infection source* | xx (xx) | xx (xx) | xx (xx) | xx (xx) |
| Dialysis – n (%) | xx (xx) | xx (xx) | xx (xx) | xx (xx) |
| **On admission to the ward** |  |  |  |  |
| Vital signs - Median (Q1, Q3) |  |  |  |  |
| Systolic blood pressure | xx (xx, xx) | xx (xx, xx) | xx (xx, xx) | xx (xx, xx) |
| Diastolic blood pressure | xx (xx, xx) | xx (xx, xx) | xx (xx, xx) | xx (xx, xx) |
| Heart rate (beats/minute) | xx (xx, xx) | xx (xx, xx) | xx (xx, xx) | xx (xx, xx) |
| Temperature (^ο^C) | xx (xx, xx) | xx (xx, xx) | xx (xx, xx) | xx (xx, xx) |
| Respiratory rate (breaths/minute) | xx (xx, xx) | xx (xx, xx) | xx (xx, xx) | xx (xx, xx) |
| Laboratory parameters |  |  |  |  |
| Lactate (mmol/L) - Median (Q1, Q3) | xx (xx, xx) | xx (xx, xx) | xx (xx, xx) | xx (xx, xx) |
| White blood cells (10^9^/L) - Median (Q1, Q3) | xx (xx, xx) | xx (xx, xx) | xx (xx, xx) | xx (xx, xx) |
| Bilirubin (µmol/L) - Median (Q1, Q3) | xx (xx, xx) | xx (xx, xx) | xx (xx, xx) | xx (xx, xx) |
| Creatinine (µmol/L) - Median (Q1, Q3) | xx (xx, xx) | xx (xx, xx) | xx (xx, xx) | xx (xx, xx) |
| Blood culture – n (%) | xx (xx) | xx (xx) | xx (xx) | xx (xx) |
| Respiratory culture – n (%) | xx (xx) | xx (xx) | xx (xx) | xx (xx) |
| Urine culture – n (%) | xx (xx) | xx (xx) | xx (xx) | xx (xx) |
| Body fluid culture (pleural, ascitic, CSF, joint) – n (%) | xx (xx) | xx (xx) | xx (xx) | xx (xx) |
| Treatments – n (%) |  |  |  |  |
| Intravenous fluids | xx (xx) | xx (xx) | xx (xx) | xx (xx) |
| Antibiotics | xx (xx) | xx (xx) | xx (xx) | xx (xx) |

***** no documented infection or infection source is defined as no ICD-10AU for pneumonia, urinary tract infection, skin and soft tissue infection, intra-abdominal infection or other infections.

**Table S2:** Alert information and process measures.

|  | **Intervention (XX)** | **Control (XX)** | **p-value** |
| --- | --- | --- | --- |
| Alert count per patient- Median (Q1, Q3) | xx (xx, xx) | xx (xx, xx) | 0.xx |
| Time to first alert - Median (Q1, Q3) | xx (xx, xx) | xx (xx, xx) | 0.xx |
| Alert information criteria leading to triggering – n (%) |  |  |  |
| Respiratory rate ≥22 (breaths/minute) | xx (xx) | xx (xx) | 0.xx |
| Systolic blood pressure ≤100 mm Hg | xx (xx) | xx (xx) | 0.xx |
| Glasgow Coma scale <15 | xx (xx) | xx (xx) | 0.xx |
| Alert acknowledgment by nurse |  |  |  |
| Percent acknowledged – n (%) | xx (xx) | - | - |
| Time to acknowledgment |  | - | - |
| 0-15 minutes – n (%) | xx (xx, xx) | - | - |
| 16-60 minutes – n (%) | xx (xx, xx) | - | - |
| >60 minutes – n (%) | xx (xx, xx) | - | - |
| Alert acknowledgment by physician |  |  |  |
| Percent acknowledged – n (%) | xx (xx) | - | - |
| Time to acknowledgment |  | - | - |
| 0-30 minutes – n (%) | xx (xx, xx) | - | - |
| 30-60 minutes – n (%) | xx (xx, xx) | - | - |
| >120 minutes – n (%) | xx (xx, xx) | - | - |
| Vital signs in the 12 hours pre-alert- Median (Q1, Q3) |  |  |  |
| Blood Pressure (mm Hg) |  |  |  |
| Systolic blood pressure | xx (xx, xx) | xx (xx, xx) | 0.xx |
| Diastolic blood pressure | xx (xx, xx) | xx (xx, xx) | 0.xx |
| Heart Rate (beats/minute) | xx (xx, xx) | xx (xx, xx) | 0.xx |
| Temperature (°C) | xx (xx, xx) | xx (xx, xx) | 0.xx |
| Respiratory Rate (breaths/minute) | xx (xx, xx) | xx (xx, xx) | 0.xx |
| Laboratory parameters in the 12 hours pre-alert |  |  |  |
| Lactate (mmol/L) - Median (Q1, Q3) | xx (xx, xx) | xx (xx, xx) | 0.xx |
| White blood cells (10^9^/L) - Median (Q1, Q3) | xx (xx, xx) | xx (xx, xx) | 0.xx |
| Bilirubin (µmol/L) - Median (Q1, Q3) | xx (xx, xx) | xx (xx, xx) | 0.xx |
| Creatinine (µmol/L) - Median (Q1, Q3) | xx (xx, xx) | xx (xx, xx) | 0.xx |
| Blood culture – n (%) | xx (xx) | xx (xx) | 0.xx |
| Respiratory culture – n (%) | xx (xx) | xx (xx) | 0.xx |
| Urine culture – n (%) | xx (xx) | xx (xx) | 0.xx |
| Body fluid culture (pleural, ascitic, CSF, joint) – n (%) | xx (xx) | xx (xx) | 0.xx |
| Treatments in the 12 hours pre-alert – n (%) |  |  |  |
| Intravenous fluids | xx (xx) | xx (xx) | 0.xx |
| Antibiotics | xx (xx) | xx (xx) | 0.xx |
| Process measures |  |  |  |
| Lactate reported in the 12 hours after the alert (n, %)* | xx (xx) | xx (xx) | 0.xx |
| Lactate highest value reported in the 12 hours after the alert* | xx (xx, xx) | xx (xx, xx) | 0.xx |
| Blood culture ordered in the 12 hours after alert (n, %)* | xx (xx) | xx (xx) | 0.xx |
| Respiratory culture ordered in the 12 hours after alert (n, %)* | xx (xx) | xx (xx) | 0.xx |
| Urine culture ordered in the 12 hours after alert (n, %)* | xx (xx) | xx (xx) | 0.xx |
| Other body fluid culture ordered in the 12 hours after alert (n, %)* | xx (xx) | xx (xx) | 0.xx |
| Intravenous fluid administered in 12 hours after alert (n, %) | xx (xx) | xx (xx) | 0.xx |
| New antibiotics administered in the 12 hours after the alert (n, %) | xx (xx) | xx (xx) | 0.xx |
| New antibiotics administered in the 3 hours after the alert (n, %) | xx (xx) | xx (xx) | 0.xx |
| Post-alert systolic blood pressure - lowest value in the 12 hours after the alert | xx (xx) | xx (xx) | 0.xx |
| Post-alert diastolic blood pressure - lowest value in the 12 hours after the alert | xx (xx) | xx (xx) | 0.xx |
| Post-alert heart rate - highest value in the 12 hours after the alert | xx (xx) | xx (xx) | 0.xx |
| Post-alert respiratory rate - highest value in the 12 hours after the alert | xx (xx) | xx (xx) | 0.xx |

If the test was not performed within the 12 hours before the alert.

**Table S3:** Outcomes.

|  | **Intention-to-treat cohort** | | | | | **Alert cohort** | | | | | |
| --- | --- | --- | --- | --- | --- | --- | --- | --- | --- | --- | --- |
|  | **Intervention (XX)** | **Control (XX)** | **RR**  **(95% CI)** | **p-value** | **FDR** | **Intervention (XX)** | **Control (XX)** | **p-value**  **(XX)** | **RR**  **(95% CI)** | **p-value** | **FDR** |
| 90-day in-hospital mortality-n (%) | xx (xx) | xx (xx) | xx.x (xx.x, xx.x) | 0.xx | 0.xx | xx (xx) | xx (xx) | 0.xx | xx.x (xx.x, xx.x) | 0.xx | 0.xx |
| ICU admission – n (%) | xx (xx) | xx (xx) | xx.x (xx.x, xx.x) | 0.xx | 0.xx | xx (xx) | xx (xx) | 0.xx | xx.x (xx.x, xx.x) | 0.xx | 0.xx |
| Incident renal replacement therapy – n (%) | xx (xx) | xx (xx) | xx.x (xx.x, xx.x) | 0.xx | 0.xx | xx (xx) | xx (xx) | 0.xx | xx.x (xx.x, xx.x) | 0.xx | 0.xx |
| Vasopressor therapy – n (%) | xx (xx) | xx (xx) | xx.x (xx.x, xx.x) | 0.xx | 0.xx | xx (xx) | xx (xx) | 0.xx | xx.x (xx.x, xx.x) | 0.xx | 0.xx |
| Mechanical ventilation – n (%) | xx (xx) | xx (xx) | xx.x (xx.x, xx.x) | 0.xx | 0.xx | xx (xx) | xx (xx) | 0.xx | xx.x (xx.x, xx.x) | 0.xx | 0.xx |
| Critical Care Rapid Response Team (CCRT) activation – n (%) | xx (xx) | xx (xx) | xx.x (xx.x, xx.x) | 0.xx | 0.xx | xx (xx) | xx (xx) | 0.xx | xx.x (xx.x, xx.x) | 0.xx | 0.xx |
| Cardiac arrest – n (%) | xx (xx) | xx (xx) | xx.x (xx.x, xx.x) | 0.xx | 0.xx | xx (xx) | xx (xx) | 0.xx | xx.x (xx.x, xx.x) | 0.xx | 0.xx |
| Multidrug-resistant organism – n (%) | xx (xx) | xx (xx) | xx.x (xx.x, xx.x) | 0.xx | 0.xx | xx (xx) | xx (xx) | 0.xx | xx.x (xx.x, xx.x) | 0.xx | 0.xx |
| Clostridium difficile infection – n (%) | xx (xx) | xx (xx) | xx.x (xx.x, xx.x) | 0.xx | 0.xx | xx (xx) | xx (xx) | 0.xx | xx.x (xx.x, xx.x) | 0.xx | 0.xx |
| Continuous outcomes |  |  | Beta estimates (95% CI) |  |  |  |  |  |  |  |  |
| Hospital length of stay (days), median (Q1, Q3) | xx (xx, xx) | xx (xx, xx) | xx.x (xx.x, xx.x) | 0.xx | 0.xx | xx (xx, xx) | xx (xx, xx) | 0.xx | xx.x (xx.x, xx.x) | 0.xx | 0.xx |
| ICU-free days, median (Q1, Q3) | xx (xx, xx) | xx (xx, xx) | xx.x (xx.x, xx.x) | 0.xx | 0.xx | xx (xx, xx) | xx (xx, xx) | 0.xx | xx.x (xx.x, xx.x) | 0.xx | 0.xx |
| Antibiotic free-days up to 90 days, median (Q1, Q3) | xx (xx, xx) | xx (xx, xx) | xx.x (xx.x, xx.x) | 0.xx | 0.xx | xx (xx, xx) | xx (xx, xx) | 0.xx | xx.x (xx.x, xx.x) | 0.xx | 0.xx |

CI: confidence interval, FDR: false discovery rate, ICU: intensive care unit, Q1: first quartile, Q3: third quartile, RR: relative risk

**Table S4:** Post-alert outcomes in the alert cohort.

|  | **Alert cohort** | | | | | |
| --- | --- | --- | --- | --- | --- | --- |
|  | **Intervention (XX)** | **Control**  **(XX)** | **p-value**  **(XX)** | **RR**  **(95% CI)** | **p-value** | **FDR** |
| Post-alert ICU admission within 14 days – n (%) | xx (xx) | xx (xx) | 0.xx | xx.x (xx.x, xx.x) | 0.xx | 0.xx |
| Post-alert renal replacement therapy (RRT) within 14 days – n (%) | xx (xx) | xx (xx) | 0.xx | xx.x (xx.x, xx.x) | 0.xx | 0.xx |
| Post-alert vasopressors within 14 days – n (%) | xx (xx) | xx (xx) | 0.xx | xx.x (xx.x, xx.x) | 0.xx | 0.xx |
| Post-alert Mechanical ventilation within 14 days – n (%) | xx (xx) | xx (xx) | 0.xx | xx.x (xx.x, xx.x) | 0.xx | 0.xx |
| Post-alert CCRT activation within 14 days – n (%) | xx (xx) | xx (xx) | 0.xx | xx.x (xx.x, xx.x) | 0.xx | 0.xx |
| Post-alert code blue within 14 days – n (%) | xx (xx) | xx (xx) | 0.xx | xx.x (xx.x, xx.x) | 0.xx | 0.xx |
| Post-alert antibiotics within 14 days – n (%) | xx (xx) | xx (xx) | 0.xx | xx.x (xx.x, xx.x) | 0.xx | 0.xx |
| Post-alert MDROs up to day 90 – n (%) | xx (xx) | xx (xx) | 0.xx | xx.x (xx.x, xx.x) | 0.xx | 0.xx |
| Post Alert Clostridium difficile infection up to day 90 – n (%) | xx (xx) | xx (xx) | 0.xx | xx.x (xx.x, xx.x) | 0.xx | 0.xx |

CI: confidence interval, ICU: intensive care unit, CCRT: critical care response team, MDRO: multidrug drug resistant organism

**Table S5:** Subgroup Analysis; ITT population.

|  | **Intervention (XX)** | **Control (XX)** | **p-value (XX)** | **RR (95% CI)** | **p-value** | **Interaction p** | **FDR** |
| --- | --- | --- | --- | --- | --- | --- | --- |
| Age=< 65 years | xx (xx) | xx (xx) | 0.xx | xx.x (xx.x, xx.x) | 0.xx | 0.xx | 0.xx |
| Age >65 years | xx (xx) | xx (xx) | 0.xx | xx.x (xx.x, xx.x) | 0.xx | 0.xx | 0.xx |
| Patients with documented infection source* | xx (xx) | xx (xx) | 0.xx | xx.x (xx.x, xx.x) | 0.xx | 0.xx | 0.xx |
| Patients with no documented infection or infection source | xx (xx) | xx (xx) | 0.xx | xx.x (xx.x, xx.x) | 0.xx | 0.xx | 0.xx |
| Admitted to a medical ward | xx (xx) | xx (xx) | 0.xx | xx.x (xx.x, xx.x) | 0.xx | 0.xx | 0.xx |
| Admitted to a surgical ward | xx (xx) | xx (xx) | 0.xx | xx.x (xx.x, xx.x) | 0.xx | 0.xx | 0.xx |
| Admitted to an oncology ward | xx (xx) | xx (xx) | 0.xx | xx.x (xx.x, xx.x) | 0.xx | 0.xx | 0.xx |
| Admitted to a mixed ward |  |  |  |  |  |  |  |
| Alert within 48 hours of admission | xx (xx) | xx (xx) | 0.xx | xx.x (xx.x, xx.x) | 0.xx | 0.xx | 0.xx |
| Alert after 48 hours of admission | xx (xx) | xx (xx) | 0.xx | xx.x (xx.x, xx.x) | 0.xx | 0.xx | 0.xx |
| Admitted to a COVID-19 ward | xx (xx) | xx (xx) | 0.xx | xx.x (xx.x, xx.x) | 0.xx | 0.xx | 0.xx |
| Admitted to a non-COVID-19 ward | xx (xx) | xx (xx) | 0.xx | xx.x (xx.x, xx.x) | 0.xx | 0.xx | 0.xx |

CI: confidence interval, RR: relative risk, FDR: false discovery rate

*documented infection source includes ICD-10AU for pneumonia, urinary tract infection, skin and soft tissue infection, intra-abdominal infection or other infections.
